# Routine germline genetic testing in 3552 unselected NHS breast cancer patients: Evidence informing testing criteria and implementation of a ‘BRCA-DIRECT’ mainstreaming pathway

**DOI:** 10.64898/2026.02.02.26344266

**Authors:** Bethany Torr, Lea Mansour, Caitlin T. Fierheller, Monica Hamill, Joshua Nolan, Nicola Bell, Subin Choi, Sophie Allen, Sudeekshna Muralidharan, Suzanne MacMahon, Yasmin Clinch, Mikel Valganon-Petrizan, Helena Harder, Alice Garrett, D. Gareth Evans, Angela George, Valerie Jenkins, Lesley Fallowfield, Rosa Legood, Zoe Kemp, Ranjit Manchanda, Clare Turnbull

## Abstract

**Background:** Breast cancer susceptibility gene testing (BCSG-testing) is expanding in relation to both eligibility for testing and number of genes included on testing panels. However, uncertainty remains regarding the most effective testing strategies for identifying clinically actionable germline pathogenic variants (gPVs) while balancing increased burden on breast and genetics clinical services.

**Patients and Methods:** The North Thames Mainstreaming of Breast Cancer Genetic Testing (NT-MBGT) programme piloted unselected breast cancer (BC) patient BCSG-testing via a clinician-light ‘BRCA-DIRECT’ mainstreaming pathway. We present ‘real-world’ evaluation of (i) gPV pick-up rates according to BC characteristics and (ii) operational feasibility, acceptability, and satisfaction with the ‘BRCA-DIRECT’ expanded testing pathway.

**Results:** The ‘BRCA-DIRECT’ pathway successfully tested 3,517 newly-diagnosed BC patients within 14 National Health Service (NHS) breast oncology units, with high levels of patient and breast healthcare professional (HCP) satisfaction, and genetics HCPs reporting concomitant decrease in service referrals.

The overall pick-up rate of gPVs was 4.7%. Current NHS eligibility criteria would have offered testing to 20.6% of patients and identified 49.2% of observed gPVs in high penetrance (HP)-BCSGs (*BRCA1*/*BRCA2*/*PALB2*) and 18.2% of gPVs in intermediate penetrance (IP)-BCSGs (*CHEK2/ATM/RAD51C/RAD51D*). ‘Ultra-simple’ eligibility criteria could improve detection (sensitivity) to 74.6% and 61.4%, respectively, whilst increasing testing to 50.2% of BC cases.

**Conclusions:** Evidence from the NT-MBGT programme demonstrates that expanding BCSG-testing via a clinician-light pathway is acceptable and feasible, without increasing the burden on limited breast and genetics workforce, and has high satisfaction. Simplified testing criteria could improve identification of gPVs in HP-BCSGs. The concomitant increased pick-up of gPVs in IP-BCSGs warrants further consideration.

**highlights:**

- In this real-world evaluation we observed the successful rollout of the ‘BRCA-DIRECT’ streamlined, clinician-light mainstreaming pathway for a pilot of germline breast cancer susceptibility gene testing in 3517 unselected breast cancer patients from 14 regional breast oncology/surgical units.
- Patients undergoing testing via the pathway reported high levels of satisfaction and low decisional regret, with breast and genetics healthcare professionals highly recommending the pathway for mainstream testing.
- Differences were observed between breast healthcare professionals preferring unselected breast cancer patient testing and genetics healthcare professionals preferring restriction to current national testing criteria due to broader concerns around equity of access to testing.
- We identified that current national testing criteria would have missed identifying 50.8% of germline pathogenic variants in high-penetrance, clinically actionable genes, likely having implications for treatment and surgical decision-making in the breast cancer patients.
- We evaluated the performance of two additional approaches for establishing testing eligibility criteria to understand how we could best balance maximising identification of germline pathogenic variants (sensitivity) whilst limiting (unnecessary) testing within the breast cancer patient population (specificity).

## Introduction

Clinical breast cancer susceptibility gene testing (BCSG-testing) has been performed since the late 1990s following identification of *BRCA1* and *BRCA2*. Historically, testing was typically offered to a small number of individuals with a very strong personal and family history of hereditary breast/ovarian cancer (HBOC). Testing usually took many months, limiting applicability for immediate surgical or oncological management of the proband. BCSG-testing was often initiated by concerned family members rather than clinicians managing breast cancer (BC) patients. Identification of a germline pathogenic variant (gPV) enabled predictive testing to dichotomise family members as at very-high-risk of, or spared from, the inheritable disease.

Advances in genetic technologies, in particular Next Generation Sequencing, have dramatically progressed the scale, speed, cost and accuracy of genetic testing. Enabling high-throughput, higher-volume testing of many genes at relatively affordable rates, deliverable within weeks (or even days). This has transformed delivery of BCSG-testing.

Firstly, eligibility criteria have been extended. In many healthcare systems, a sizeable proportion of BC patients may now be eligible for BCSG-testing, based solely on their own BC (e.g. young onset disease, triple negative histology or bilateral). Other cases of female BC are now eligible based on a comparatively modest family history. ^1^

Secondly, the number of genes tested has been expanded. Until recently, testing included *BRCA1*/*BRCA2*, based on clear-cut actionability for concomitant high risks of BC and ovarian cancer (OC). ^2^ Testing now typically includes additional BCSGs, delivered as a multi-gene panel test (MGPT).^3^ BCSGs such as *PALB2*, *CHEK2, ATM, RAD51C, RAD51D* and *BARD1* were identified in case control studies of candidate genes, selected based on similarities to *BRCA1/BRCA2* for their roles in DNA repair pathways. These genes have lower penetrance for BC than *BRCA1/BRCA2*. Additionally, MGPT may include very rare pleomorphic cancer predisposition syndrome genes, such as *TP53, STK11, PTEN, NF1* and *CDH1*, which involve BC susceptibility in combination with a constellation of rare cancers and/or characteristic non-malignant phenotypic features. ^4^

Thirdly, rather than referral into clinical genetics, testing is increasingly delivered by mainstream oncology clinicians alongside other diagnostic investigations. Opportunities for surgical decision-making and advances in systemic oncological management, in particular PARP inhibitors (PARPi), along with logistic feasibility, have driven rapid testing earlier in the pathway. ‘Mainstreaming’ of higher volumes of BC patients has necessitated pathway re-design.^5–7^ We have previously reported our ‘BRCA-DIRECT’ pathway providing digitally-supported, clinician-light BCSG-testing using home saliva testing supported by a genetic counsellor hotline, within a randomised trial at five National Health Service (NHS) oncology centres comprising >1000 BC patients. ^8^

In the United Kingdom (UK), NHS BCSG-testing comprises seven genes and ∼20% of BC cases meet NHS criteria (with comparable proportions eligible in other European healthcare services).^9^ These selection criteria, relating to BC characteristics and family history of relevant cancers, enrich for gPVs and improve the detection rate per-test. For example, testing 20% of BCs (using UK NHS criteria) has been estimated to pick-up ∼50% of BC cases with *BRCA1/BRCA2* gPVs. ^9^ However: (i) half or more gPVs are missed, (ii) family history assessment can be complex and laborious and (iii) there is messy overlap between eligibility based on personal/family history and PARPi therapeutic eligibility. ^10, 11^

In the broader context of healthcare systems placing increasing focus on screening, prevention and early diagnosis (SPED), these factors are driving calls for expansion of germline genetic testing in BC cases, simplifying eligibility criteria, and improving sensitivity in identifying gPVs. ^11–15^ However, several questions remain unaddressed:

i. What proportion of BC cases should receive BCSG-testing; can we expand and simplify eligibility?
ii. Which genes should we test? Whilst SPED interventions for *BRCA1/BRCA2* are well-evaluated, the impacts and benefits of detecting PVs in the other genes are less well-established.
iii. How best can testing be delivered safely, robustly, and within tight time-scales to larger numbers of women through busy BC clinics, whilst ensuring well-informed consent and necessary psycho-social support?

We aimed to address these questions under ‘real-world conditions’ via the North Thames Genomics Laboratory Hub Mainstreaming of Breast Cancer Genetics Programme (NT-MBCG).

## Methods

Within the NT-MBCG programme, a ‘BRCA-DIRECT’ pathway was piloted for BCSG-testing at 14 NHS hospitals across greater-London, UK. ^16^ Including teaching hospitals and smaller district hospitals, referring into the North Thames Genomics Laboratory Hub for NHS cancer genetic testing. Each hospital had a named Clinical Nurse Specialist ‘champion’ and Clinical lead to oversee implementation and delivery.

### Patients

#### Inclusion Criteria

- Female,
- >18-years, and
- within 12-months of a new presentation of high-grade ductal carcinoma in-situ (DCIS) or invasive BC (including a new primary, bilateral, recurrent or metastatic disease).

This extended beyond NHS criteria (Table 1), enabling prospective BCSG-testing for all patients with a new presentation of BC (unselected).

**Table 1.**
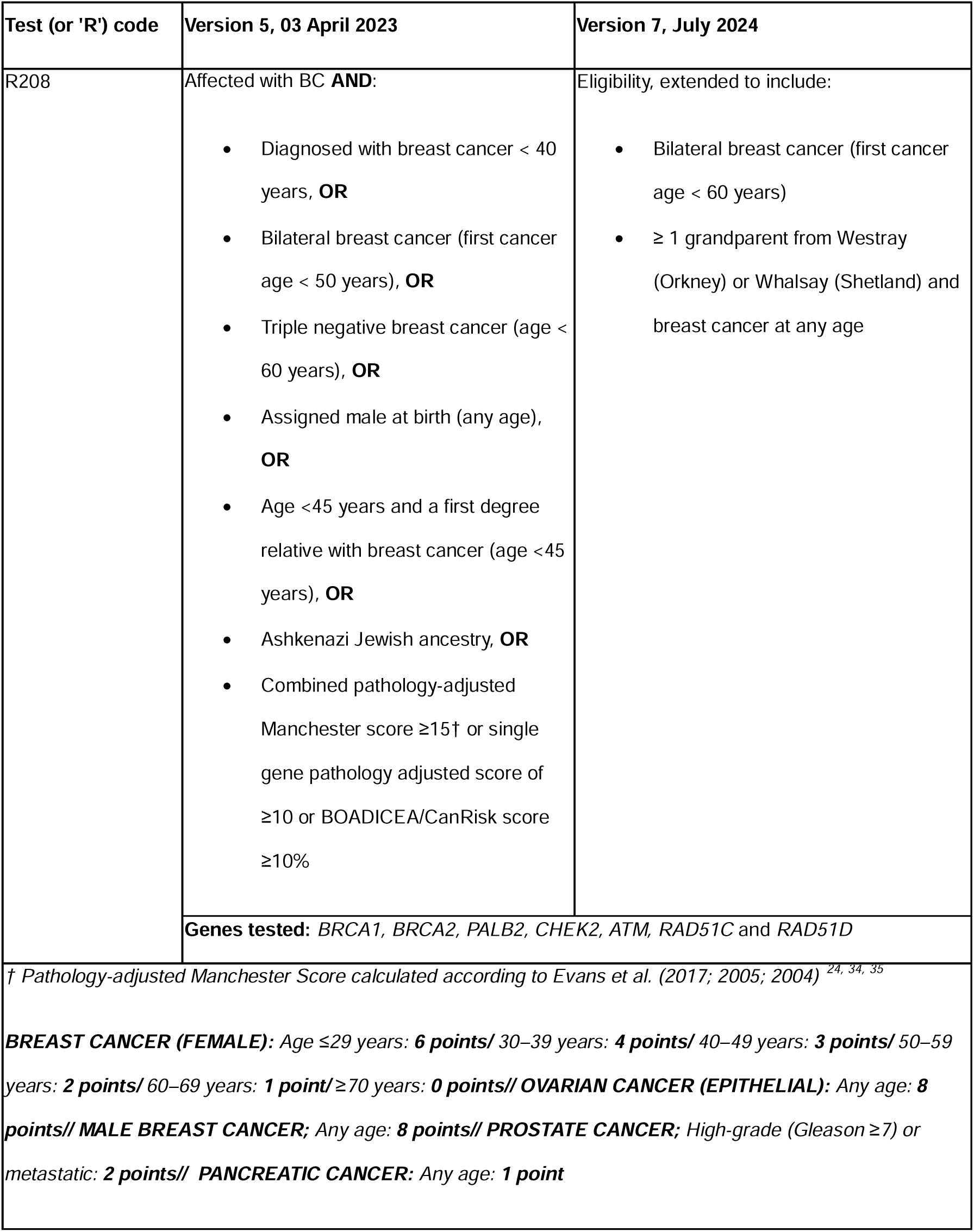
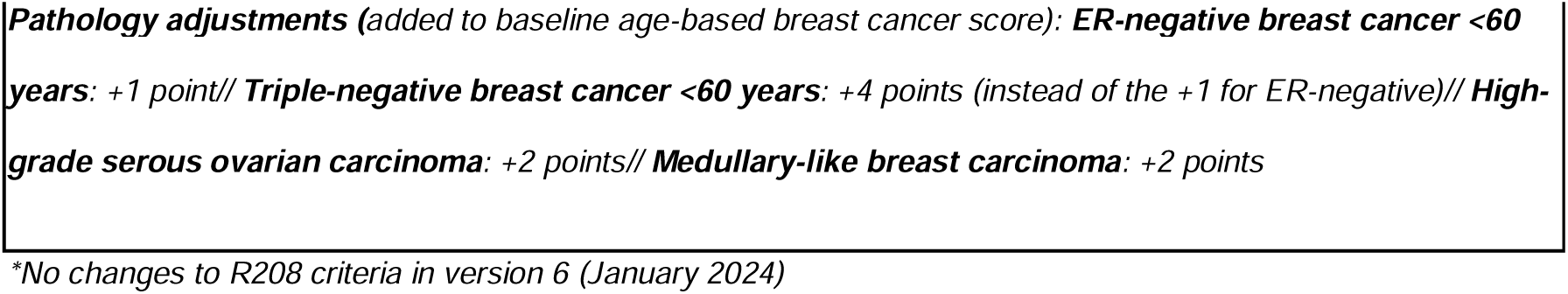
National Genetic Testing Criteria. As extracted from the National Genomic Test Directory for Rare and Inherited Diseases in England ^17^

**Table 2.**
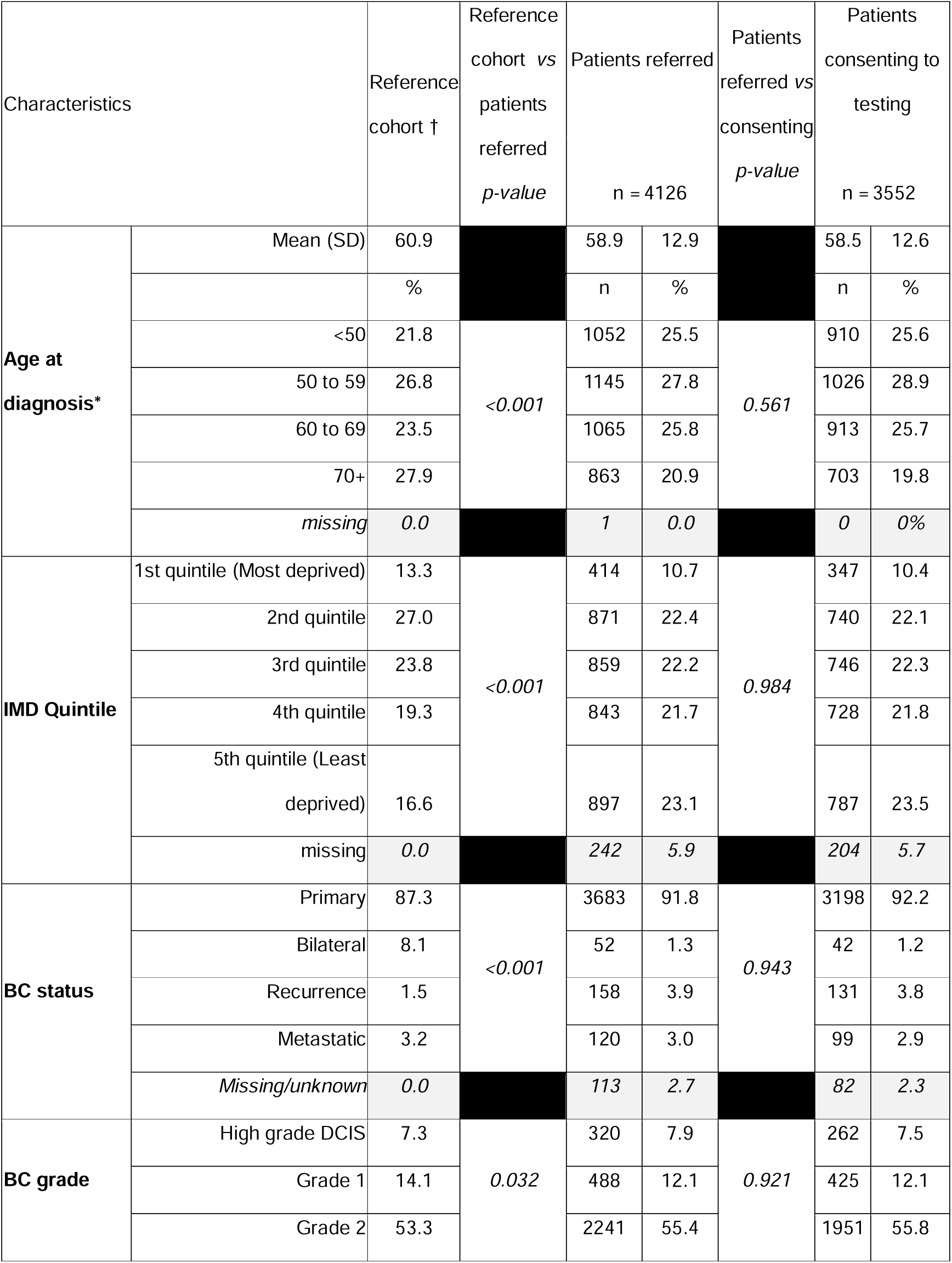
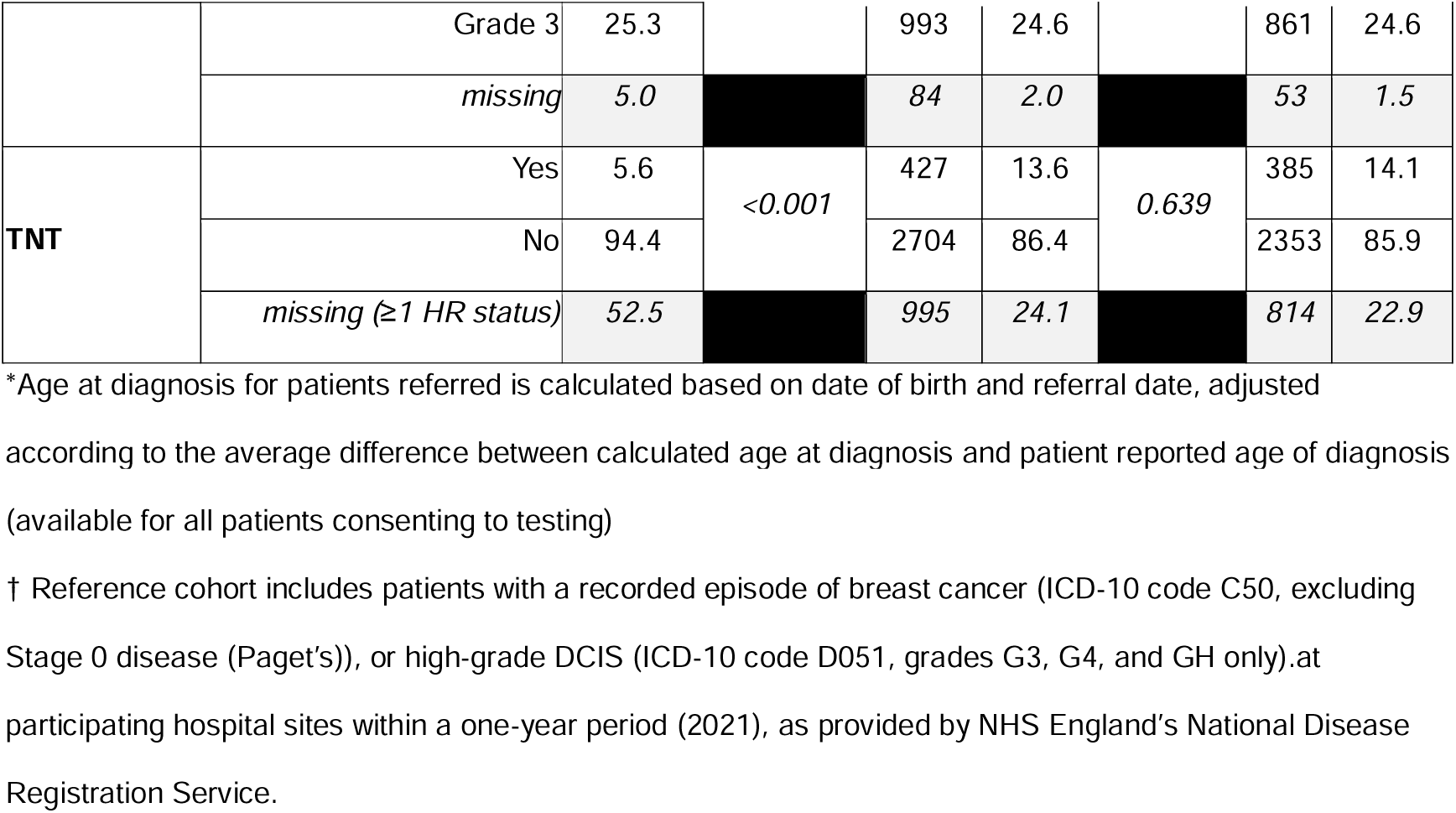
Characteristics of patients. . Including a comparison of all patients referred and consenting to genetic testing via the NT-MBCG programme compared with the baseline characteristics of all breast cancer cases diagnosed.

#### Exclusion Criteria

- Patients who previously had NHS BCSG-testing.

Male BC patients and those eligible for *TP53* testing (BC ≤30 years; HER2 positive BC ≤35 years) were referred for testing at regional clinical genetics services (RCGSs).

#### Pathway

Patients were offered BCSG-testing via BRCA-DIRECT by their breast oncology or surgical teams. Local models of the BRCA-DIRECT pathway varied minorly given local needs and practices; however, the following pathway principles remained the same.

#### Pre-test counselling and consent

Interested patients were provided with a BRCA-DIRECT testing pack, including: a testing information booklet, personal details and consent form, and saliva sampling kit. Additional information about the testing process, including videos and translated versions of material, were accessible online (QR-code and URL within the information booklet). The testing pack could be completed in clinic or at-home and returned using freepost packaging provided to the RCGS at the Royal Marsden NHS Foundation Trust (RMH)/Institute of Cancer Research.

Genetic Nurse/Counsellor (GN/C) support was available throughout the pathway via a telephone helpline (available 9am-to-5pm, Monday-to-Friday, with extended hours on Wednesday). Translators and video calling with subtitling were available, if required. Administrative support was available via telephone helpline or email.

#### Return of results and downstream management

Test results were returned via post to the patient and their general practitioner and emailed to the referring breast oncology/surgical team. Patients with a gPV or variant of uncertain significance (VUS) were offered a follow-up appointment with a BRCA-DIRECT GN/C to discuss the results and cascade testing within one week of posting the result. Positive patients were then referred to their RCGS for downstream management, and the Very-High-Risk NHS breast screening service (where appropriate).

#### Genetic Testing

The primary method of DNA sampling utilised saliva samples (self-collected by patients). Extraction and sequencing were performed within a single NHS accredited laboratory (RMH). Full sequence analysis of all coding exons for small variants in seven genes (*BRCA1/BRCA2/PALB2/CHEK2/ATM/RAD51C*/*RAD51D*) was performed, according to NHS England’s National Genomic Test Directory for Rare and Inherited Diseases, criteria ‘R208’.^17^ Where next generation sequencing analysis suggested a copy number variant (CNV), multiplex ligation-dependent probe amplification was also performed.

Variants were classified in accordance with guidelines from the American College of Medical Genetics and Genomics and the Association for Molecular Pathology and Cancer Variant Interpretation Group UK.^18–21^ As per UK Association of Clinical Genomic Scientists recommendations, only variants classified as pathogenic, likely pathogenic or ‘hot’ VUS (where there is a significant chance of the variant being upgraded to [likely] pathogenic) were reported. Only protein truncating variants were reported for *CHEK2, ATM, RAD51C* and *RAD51D* (in addition to NM_000051.4(ATM):c.7271T>G (p.Val2424Gly)).

#### Objectives

The primary objective was to assess pickup rates of gPVs across seven genes on the R208 panel, to inform evaluation regarding expansions of current testing eligibility criteria.

Secondary objectives (SO) related to acceptance, feasibility, and satisfaction with the BRCA-DIRECT pathway for mainstream BCSG-testing:

**SO1.** Evaluate uptake of genetic testing via the BRCA-DIRECT mainstreaming pathway by different patients across 14 breast oncology settings.

**SO2.** Assess performance of saliva sampling as the primary DNA sampling method.

**SO3.** Understand clinician experience of delivering the BRCA-DIRECT pathway and preferences for future rollout.

**SO4.** Evaluate patient satisfaction and decisional regret for those undergoing testing via BRCA-DIRECT.

### Data collection and analysis

#### Baseline data collection

Age, BC status, and tumour pathology were provided by clinical teams. Personal and family history of cancer, and ancestry were collected from patients. Additional demographics, including index of multiple deprivation (IMD) status (derived from postcode), were captured from electronic health records. Genetic testing data, sampling method and test outcomes were obtained from laboratory reports.

#### Primary outcome: pick-up rates from unselected BCSG-testing

For each patient we: (i) calculated pathology-adjusted Manchester Score (MS) and (ii) determined testing eligibility against NHS criteria at the time-point of undergoing testing (see table 1).

Data analysis was conducted within Stata version 17, with two-sided χ2 testing with Yates correction to identify significant differences between groups. We did not correct for multiple testing in our analyses.

#### SO1: Uptake of genetic testing via the BRCA-DIRECT pathway

Uptake of genetic testing was descriptively analysed for the cohort, with two-sided χ2 testing with Yates correction to identify significant differences in the distribution of demographics and BC features of patients undergoing testing via BRCA-DIRECT, compared to a reference cohort.

##### Reference cohort

Included all patients with new presentation of BC, diagnosed in a one-year period (2021) at participating hospitals. Summary level demographics and BC data were ascertained from NHS England’s National Disease Registration Service. See supplementary table 1 for details.

#### SO2 saliva sampling

Data relating to samples tested within NT-MBCG (including sampling method, outcome and failure reasons) was analysed descriptively using Stata version 17.

#### SO3: Healthcare professional experience and preferences

A digital survey (supplementary) was circulated to breast oncology and surgical healthcare professionals (HCPs) from participating hospitals via champions and to clinical genetics HCPs within RCGSs servicing these hospitals. Descriptive data analysis was conducted within Excel.

#### SO4: Patient satisfaction and decisional-regret

Following completion of testing, patients were invited to complete a validated decisional satisfaction and regret scale regarding their decision to undergo BCSG-testing via BRCA-DIRECT. ^22, 23^ Data were analysed in Stata version 17, and a Kruskal–Wallis H test evaluated differences in satisfaction scores between groups.

## Results

Between 23 June 2023 and 31 December 2024, 4126 BC patients were referred to the NT-MBCG programme for BCSG-testing (Figure 1). Of these: 43/4126 (1.0%) were ineligible for testing, 6/4126 (0.1%) were subsequently offered testing via clinical genetics, 30/4126 (0.7%) decided not to proceed with testing (active-decline), 495/4126 (12.0%) failed to return their saliva sample/consent form (passive-decline), and 3552/4126 (86.1%) consented to BCSG-testing.

**Figure 1.**
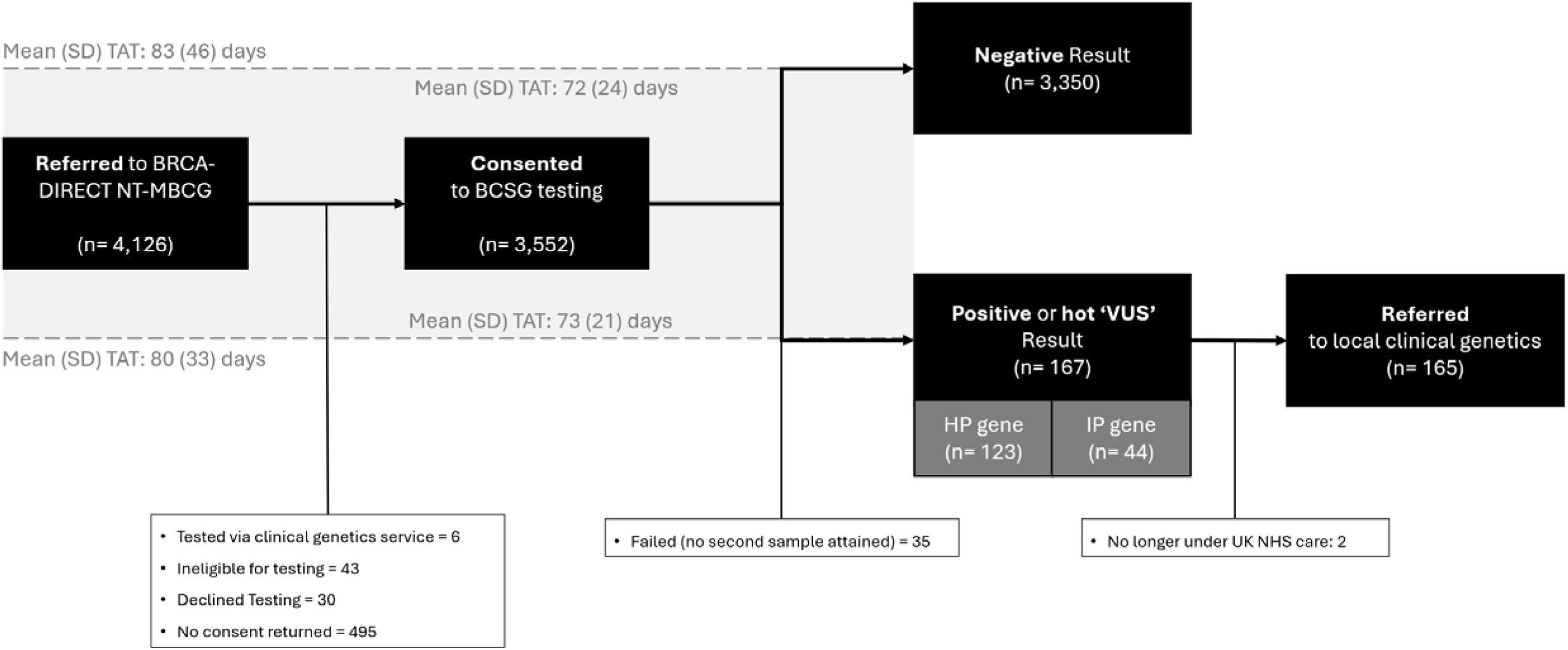
**Patient flow diagram**

### Patient demographics

For patients referred, the mean (SD) age of BC diagnosis was 58.9 (12.9) (range: 23-to-96 years) (see **Error! Reference source not found.**). 320/4126 (7.8%) had high grade DCIS, 3722/4126 (90.2%) had invasive BC; 52/4126 (1.3%) with bilateral BCs, and 427/4126 (10.3%) triple negative tumours (TNT) (**Error! Reference source not found.)**.

Compared with the reference cohort, referred patients were significantly less deprived (IMD quintile), younger in age, had primary BC, or TNT. Demographics of referred patients did not significantly differ from those consenting to testing.

### Saliva sampling and testing turnaround times

Testing was successfully completed on 3419/3552 (96.3%) primary saliva samples, with 133/3552 (3.7%) failing.

After primary sample failure, 35/133 (26.3%) patients failed to provide a second saliva sample. 98/133 (73.7%) second saliva samples were received, with 93/98 (94.9%) successfully tested and 5/98 (5.1%) failing, necessitating a blood sample. Overall saliva sample failure rate (excluding CNV fails) was 138/3650 (3.8%): 22/138 (15.9%) extraction failures, 80/138 (58.0%) sequencing fails, and 36/138 (26.1%) technical laboratory issues.

Testing was completed for 3517/3552 (99.0%) patients overall. Mean (SD) turnaround time of test results from point-of-referral was 80 (33) days for positive/VUS and 83 (46) days for negative results (Figure 1).

### Identification of pathogenic variants

Overall gPV identification rate was 166/3517 (4.7%), compromising 39/166 (23.5%) *BRCA1,* 61/166 (36.7%) *BRCA2,* 22/166 (13.3%) *PALB2,* 25/166 (15.1%) *CHEK2,* 16/166 (9.6%) *ATM,* 3/166 (1.8%) *RAD51C/D* (table 3). One *BRCA1* VUS was identified.

**Table 3.**
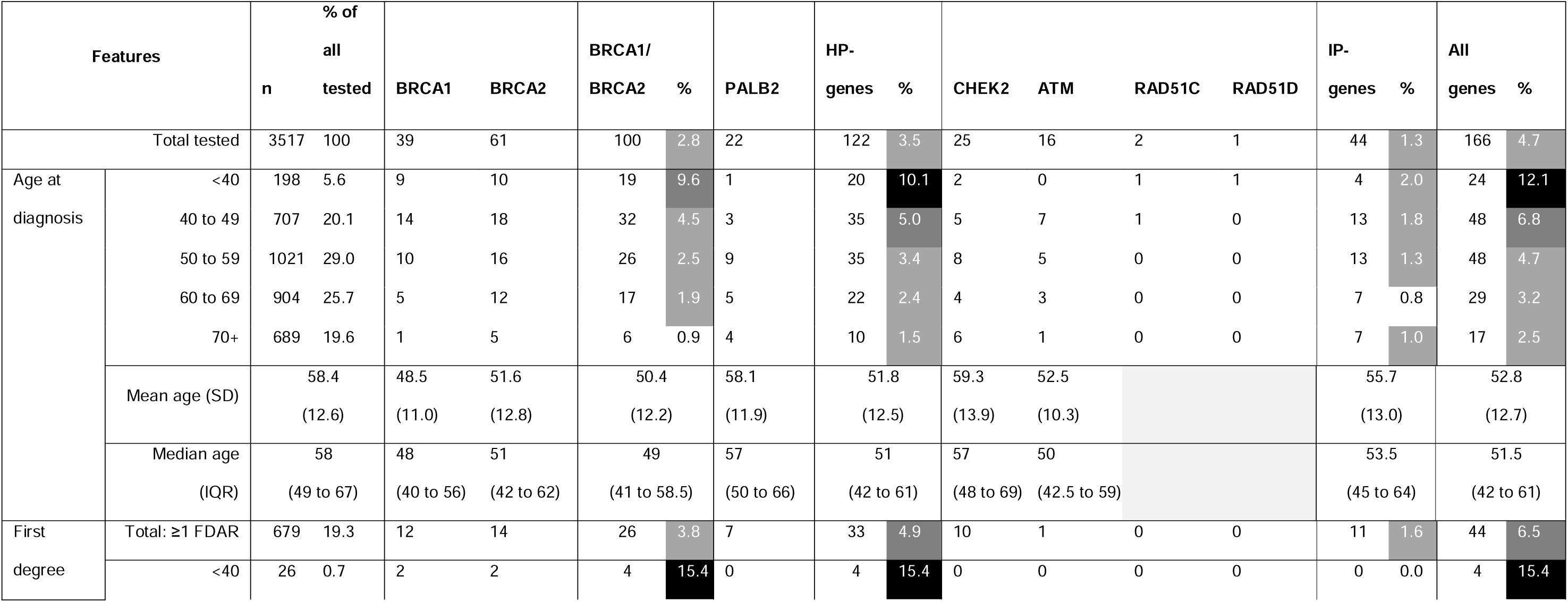
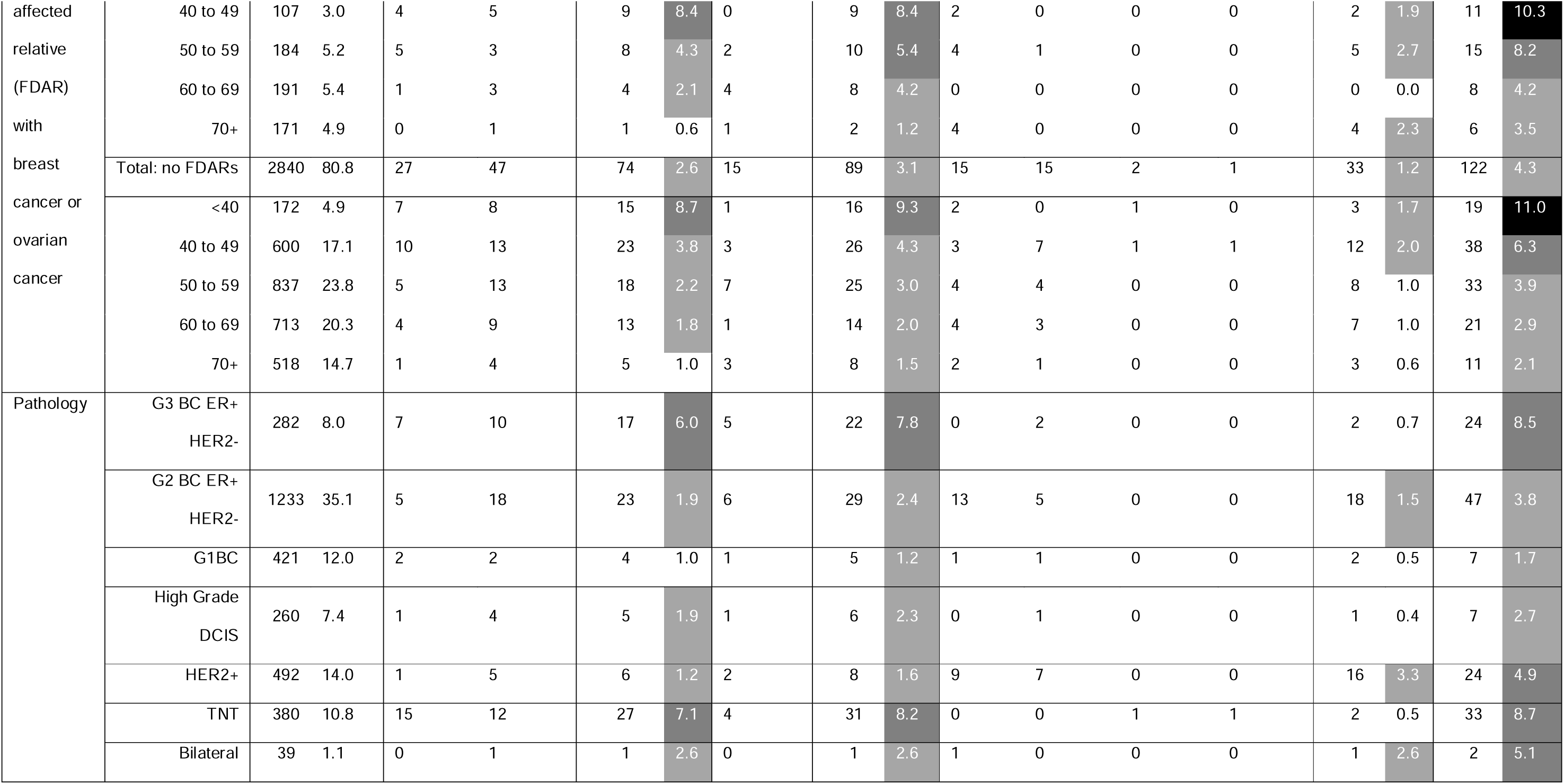
Population based testing of breast cancer by age at diagnosis, family history, and tumour pathology. Including the identification of germline pathogenic variants (**gPVs**) in high penetrance (**HP,** BRCA1, BRCA2, PALB2), intermediate penetrance (**IP,** CHEK2, ATM, RAD51C, RAD51D), and all breast cancer susceptibility genes combined. **FDAR**: First degree affected relative. **BC**: breast cancer. **G1**: Grade one invasive breast cancer. **G2**: Grade two invasive breast cancer. **G3**: grade three invasive breast cancer. **DCIS:** Ductal carcinoma in situ. **TNT**: Triple negative tumour.

### Age

The mean (SD) age of patients with gPVs was 52.8 (12.7), range 29-to-88 years (table 3). *BRCA1/BRCA2/ATM* gPV-carriers were significantly younger than the overall population (*p<0.001*).

### Tumour pathology

Where known for patients with results, 421/3464 (12.2%) patients had grade-1 disease, with HP-BCSG-gPVs identified in 5/421 (1.2%), compared with 116/3043 (3.8%) in high-grade DCIS or grade-2/3 invasive BCs. gPVs were identified in 33/380 (8.7%) TNT, 24/282 (8.5%) grade-3 BC ER+ HER2-BCs, and 2/39 (5.1%) bilateral BCs (**Table 3**).

### Testing criteria performance

We examined testing of patients based on three criteria: (i) current NHS eligibility (**Table 1**), (ii) pathology-adjusted MS at different thresholds (scores reflect the estimated probability of identifying a *BRCA1/2* gPV based on simplified tallying of personal/family cancer history), and (iii) new ‘ultra-simple’ criteria (comprising BC of age <50 or TNT or Bilateral or any first-degree BC/OC-affected relatives or AJ ancestry). We used balanced accuracy to measure performance, noting its equal weighting of sensitivity and specificity (**Table 4**).

**Table 4.**
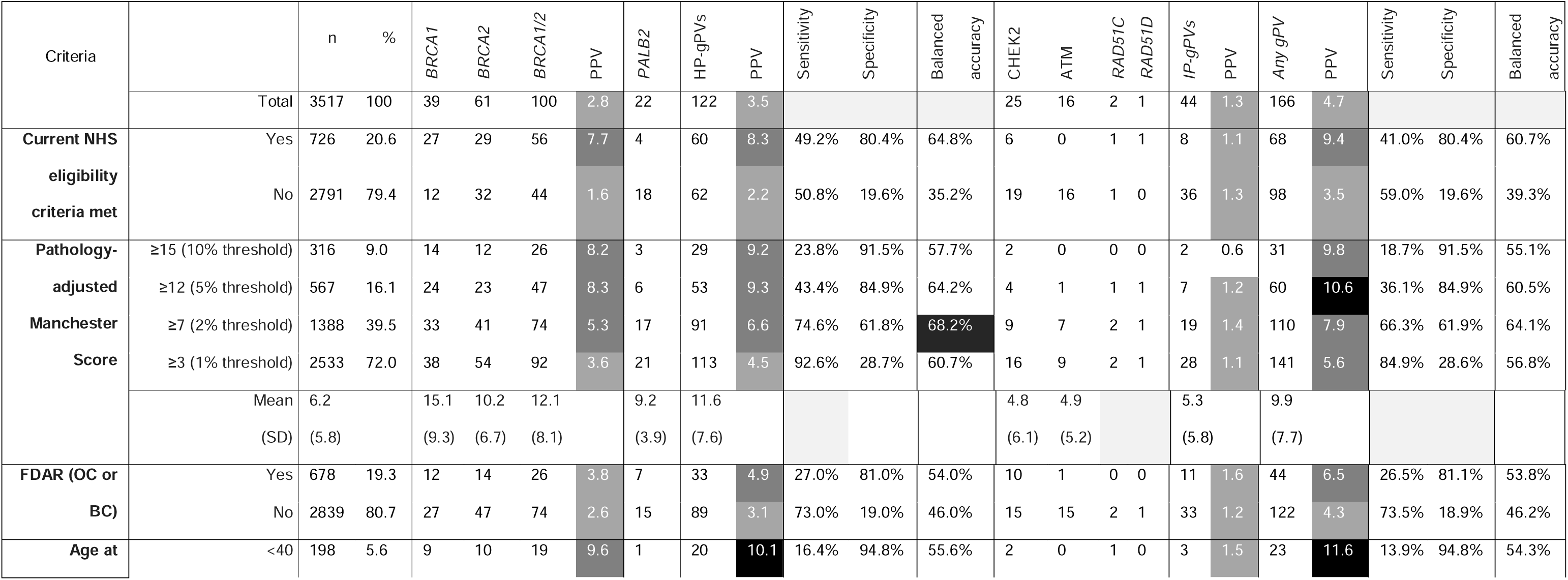
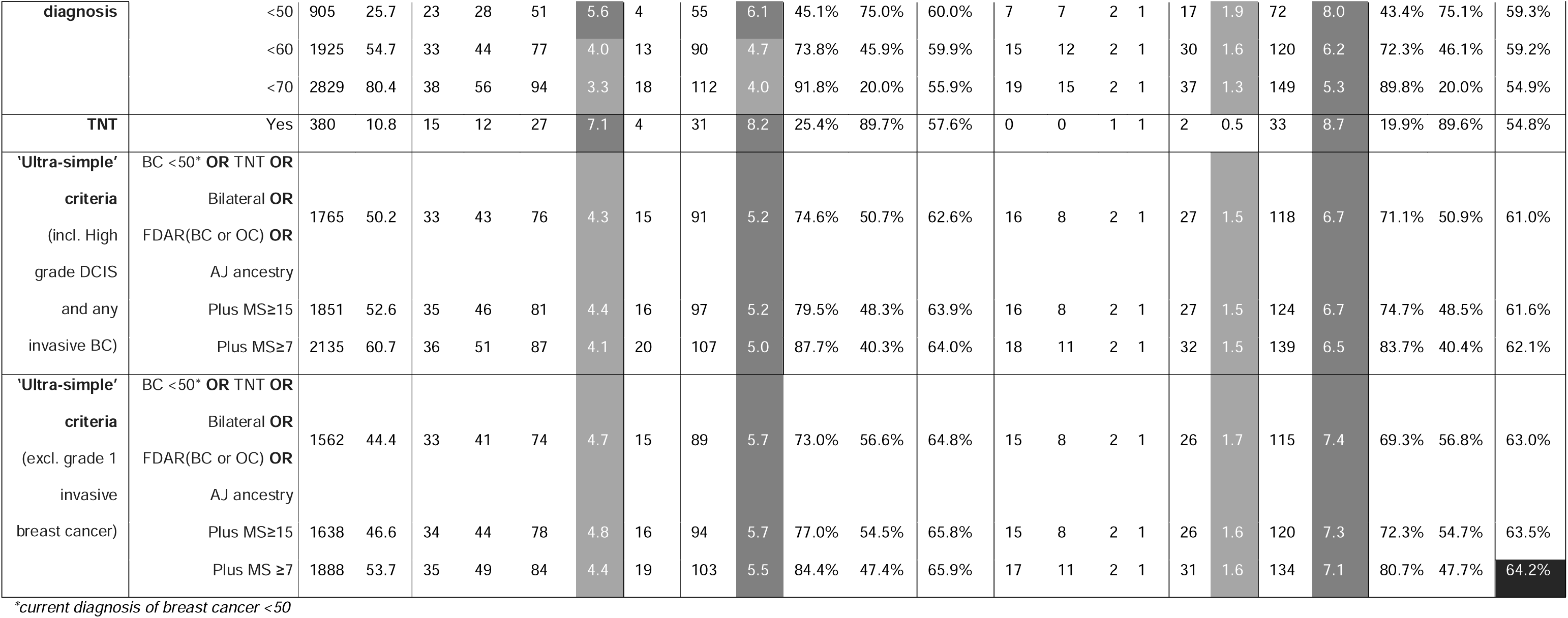
Germline pathogenic variants (gPVs) observed in high-penetrance (HP-), intermediate-penetrance (IP), and all breast cancer susceptibility genes (BCSGs) tested. . Included a breakdown based on various eligibility criteria, namely: current NHS eligibility, different thresholds of Manchester Score (**MS**), and a ‘ultra-simple’ criteria (including and excluding grade 1 breast cancers). **PPV**: Positive Predictive Value. **FDAR**: first degree affected relative with either breast cancer (**BC**) or ovarian cancer (**OC**). **AJ**: Ashkenazi Jewish ancestry. **DCIS**: Ductal carcinoma in situ.

### NHS testing criteria

NHS criteria were met in 726/3517 (20.6%) patients, identifying 68/166 (41.0%) gPVs (tables 1, 4). Overall achieving a balanced accuracy for all gPVs of 60.7% and for HP-BCSG-gPVs of 64.8%.

Proportions identified to have gPVs meeting the criteria differed significantly (P<0.001) between the HP-BCSGs and IP-BCSGs: 60/122 (49.2%) gPVs in HP-BCSGs (27/39BRCA1; 29/61 *BRCA2*; 4/22*PALB2) and* 8/44 (18.2%) gPVs in IP-BCSGs (6/25 *CHEK; 0/16 ATM; 1/2 RAD51C*; 1/1 *RAD51D)*.

### Pathology Adjusted Manchester Scoring System (PA-MSS)

The PAMSS was devised to enable simple tallying of points based on cancer pathology, age at diagnosis and family structure (table 1). The PAMS=15 threshold is equates to a ∼10% rate of detection of *BRCA1/BRCA2* gPVs, PAMS=12 to 5% and PAMS=7 to 2%. ^24^ Mean (SD) PAMS for the group overall was 6.2 (5.8) (range: - 7-to-45) (table 4). MS was strongly predictive of gPVs in HP-BCSGs, but not IP-BCSGs (supplementary table 2), with significantly higher MS in those with gPVs in HP-BCSGs (11.6 (7.6)) compared with IP-BCSGs (5.3 (5.8)) (p=0.017).

A MS>12 had similar balanced accuracy to NHS criteria for detecting HP-BCSG gPVs (64.2% vs 64.8%, respectively) with higher specificity (84.9% vs 80.4%, respectively), but lower sensitivity (43.4% vs 49.2%, respectively) (table 4). The balanced accuracy for MS>15 was lower than NHS criteria (57.7%); the high specificity (91.5%) being outweighed by much lower sensitivity (23.8%). Overall, MS>7 achieved the best balanced accuracy for HP-BCSG gPVs of all criteria tested (68.2%), reflecting improvement in detection of HP-BCSG-gPVs (sensitivity:74.6%) balanced against more modest increase in testing to 39.5% of all BC patients (specificity:61.8%).

### Ultra-Simple mainstreaming criteria

Our ‘ultra-simple’ eligibility criteria (table 4), achieved balanced accuracy for HP-BCSG gPVs of 62.6%. Additionally excluding grade-1 BCs boosted balanced accuracy to 64.8% for HP-BCSG-gPVs (similar to NHS criteria) as the reduction in sensitivity was outweighed by increased specificity (reducing testing from 50.2 to 44.4% of BC cases).

However, these ‘ultra-simple’ criteria excluded 76 patients meeting current NHS criteria with MS≥15 (in whom 5 HP-BCSG-gPVs were detected). Further addition of MSS-criteria to ‘ultra-simple’ criteria improves sensitivity for HP-BCSG-gPVs (77.0% with MS≥15 or 84.4% with MS≥7) but negates the objective of ‘ultra-simplicity’.

### Healthcare professional (HCP) experience and preferences

35 HCPs (18/35, 51.4%, breast oncology/surgical; 17/35, 48.6%, genetics) completed the experience and preferences survey.

Breast HCPs estimated prior to BRCA-DIRECT, for BCSG-testing, a median (IQR) of 10.0% (5.0 to 62.0%) patients were referred into clinical genetics, 60.0% (4.0 to 82.5%) via breast surgical mainstream pathways, and 10% (4.8 to 23.8%) via oncology mainstream pathways. Notably, 3/18 breast HCPs reported <10% of patients underwent testing through mainstreaming. Reported barriers for mainstreaming included: time constraints (11/14), clinician knowledge for providing pre-test information (7/14), knowledge of NHS criteria (7/14), language/communication barriers (3/14), and access to genetics support (2/14).

Following BRCA-DIRECT implementation, 16/18 (88.9%) breast HCPs reported offering testing to ‘most’ patients, and 10/15 (66.7%) genetics HCPs reportedly observed a decrease in referrals received.

All breast HCPs and 13/17 (76.5%) genetics HCPs reported they wanted to continue with, and recommend, BCSG-testing via the BRCA-DIRECT pathway. Most breast HCPs (13/18, 72.2%) preferred continuation with the same unselected testing criteria as piloted. Only 5/13 (38.5%) genetics HCPs indicated a preference for unselected testing, with (8/13 (61.5%)) preferring restriction to NHS criteria. Citing equity of access to genetic testing as the rationale.

Most HCPs (31/35, 88.6%) favoured using the current seven gene panel, with 4/35 (11.4%) breast HCPs favouring restriction to HP-BCSGs (*BRCA1*, *BRCA2*, *PALB2*).

### Patient satisfaction

Of the 601/2205 (27.3%) respondents, 4.2% (n=25) had gPVs/VUS results, while 95.8% tested negative. Median (IQR) satisfaction-score was 5 (5-to-5) (scores can range from 5-to-25 with lower scores reflecting higher satisfaction). Overall, 99% of patients were satisfied with their decision, 98% expressed ‘no regret’, 98.8% would make the same choice again, and 96% disagreed/strongly-disagreed that the choice did a lot of harm (Table 5). Of respondents with gPVs, 23/24 (95.8%) reported high satisfaction and ‘no regret’, while 16/24 (66.7%) disagreed/strongly-disagreed that the choice did a lot of harm. Younger patients and those from more deprived areas (1st/2nd IMD-quintiles) were relatively under-represented amongst respondents (supplementary table 3). Nevertheless, satisfaction scores did not significantly differ across IMD-quintiles (Kruskal–Wallis H-test p=0.057).

**Table 5.** Satisfaction and Regret Scale. Responses from 601/2205 (27.3%) patients sent the satisfaction and regret questionnaire, after receiving their genetic test result.

|  |  |  |  |  |  |
| --- | --- | --- | --- | --- | --- |
|  |  | I regret the | I would go for the | The choice did | The decision |
|  | It was the right | choice that | same choice if I | me a lot of | was a wise |
|  | decision | was made | had to do it over | harm | one |

|  |  |  |  |  | again |  |  |  |  |  |
| --- | --- | --- | --- | --- | --- | --- | --- | --- | --- | --- |
|  | n | % | n | % | n | % | n | % | n | % |
| **Strongly agree** | 495 | 82.36 | 1 | 0.17 | 494 | 82.2 | 1 | 0.17 | 492 | 81.86 |
| **Agree** | 102 | 16.97 | 3 | 0.5 | 100 | 16.64 | 3 | 0.5 | 100 | 16.64 |
| **Neither agree not**<br>**disagree** | 3 | 0.5 | 6 | 1 | 4 | 0.67 | 19 | 3.17 | 7 | 1.16 |
| **Disagree** | 0 | 0 | 102 | 17.03 | 1 | 0.17 | 65 | 10.85 | 0 | 0 |
| **Strongly disagree** | 1 | 0.17 | 487 | 81.3 | 2 | 0.33 | 511 | 85.31 | 2 | 0.33 |

## Discussion

We report the largest to-date prospective ‘real-world’ implementation programme of mainstream BCSG-testing delivered under standard-of-care conditions to unselected BC patients within the UK NHS, across diverse hospital settings. We evaluated, the feasibility and outcomes of unselected testing, utilising the clinician-light, oncology and clinical genetics integrated, mainstreaming ‘BRCA-DIRECT’ pathway. In this ‘real-world’ series, we systematically evaluated pick-up rates against current NHS criteria within HP- and IP-BCSGs.

### Patient pathway access and satisfaction

No significant differences were observed between those consenting and the referred cohort. It is reassuring that 86.0% of referred NT-MBCG patients underwent testing, higher than 70.9% of patients via a fully-digital pathway within our previous study, and similar to recent meta-analysis of other cancer mainstreaming studies. ^8,25^ Importantly, our pathway had high acceptability, high satisfaction, and low decisional regret. Overall, supporting implementation of the adapted pathway, with optional access to digital content and translated materials (rather than a digital-only solution).

### Feasibility and acceptability amongst clinical teams

We demonstrated successful implementation of the BRCA-DIRECT pathway across 14 diverse breast oncology centres. Seemingly, increasing the proportion of patients undergoing mainstreaming from oncology, with concomitant reduction in referrals reported by genetics HCPs.

Breast HCPs reported high satisfaction and all favoured continuation, citing patient preferences for (i) less invasive DNA sampling via saliva and (ii) easier access to testing. Genetics HCPs similarly supported pathway continuation, citing easing pressure on RCGS and shorter patient waiting times. The main difference amongst breast and genetics HCPs related to testing eligibility criteria. Breast HCPs strongly preferred simplification and ease of the unselected criteria within the programme (overcoming barriers highlighted here and in previous literature), whereas genetics HCPs were more hesitant about implementing expanded (unselected) eligibility criteria. ^26, 27^ This may reflect different perspectives, with oncology-HCPs more concerned about maximising gPVs for personalising current treatment, while genetics-HCPs reflecting on broader equity of access and efficiency of resource use.

### Pick-up rates from testing

NHS criteria were historically designed to achieve a gPV-detection rate of >10% for *BRCA1/BRCA2*. ^28^ We observed that the pick-up rate for current NHS criteria was 8.3% for HP-BCSGs and 9.4% across seven BCSGs. Despite gradually expanding over the last two decades, we found that current NHS criteria still missed 50.8% of HP-BCSGs-gPVs (consistent with other reports) as well as 82% of IP-BCSGs-gPVs.^29,9^

Our data enabled evaluation of various modifications of eligibility criteria. We sought to maintain simplicity in eligibility evaluation, whilst optimising sensitivity (pick-up of gPVs) and balancing specificity (proportion of BC cases tested). Producing balanced accuracy scores (which assume equal value of sensitivity and specificity).

Our ‘ultra-simple’ mainstreaming criteria were devised to best facilitate eligibility assessment by mainstreaming clinicians by avoiding detailed family history appraisal. These comprised (i) including all BC cases diagnosed <50 years, TNT or bilateral BC, first degree affected relative with BC/OC, or Ashkenazi Jewish ancestry and (ii) excluding all G1 BC. The ‘ultra-simple’ criteria maintained identical balanced accuracy to NHS criteria for HP-BCSG-gPVs (64.8%), improving sensitivity from 49.2% to 73.0%, whilst increasing test offer from 20.6% to 44.4% of BC patients. However, through simplifying capture of extended family history, these criteria missed a modest number of ‘strong’ families. Overall, of approaches evaluated, using MS≥7 provided best balanced accuracy for HP-BCSG-gPVs (68.2%), with testing of 39.5% of patients and detection of 74.6% of HP-BCSG-gPVs and 66.3% of all gPVs, whilst being relatively simple to tally and catching the ‘strong’ families.

Whilst there is clear rationale for exploring which criteria optimise detection of HP-BCSG-gPVs, it remains unclear what weight to place on detection of IP-BCSG-gPVs. Particularly since gPVs in *CHEK2* (and *ATM*) are relatively common, primarily associated with ER+ BC (good prognosis), and have marginal association with OC.^30–32^ The optimal eligibility parameters for implementation will be further informed by cost effectiveness analysis of the BRCA-DIRECT pathway, modelling longer-term health outcomes including mortality, with sensitivity analysis performed for different gene testing panels.

### Limitations

Despite unselected BC testing, compared with the reference cohort there appeared to be underrepresentation of those of an older age (>70 years), bilateral BC, G1 or G3 BC, and from more deprived areas. Notably, our comparator cohort included all BC patients diagnosed rather than those currently offered testing via existing pathways. Thus, findings may reflect BC patient genetic testing offer/uptake itself, rather than differences being attributable to the BRCA-DIRECT pathway. Indeed, other studies have identified that fewer older age patients are offered genetic testing by HCPs, and uptake is lower in those from more deprived socioeconomic backgrounds.^33^ Additionally, the reference cohort may not be entirely representative as data was taken from an earlier year (2021) potentially affected by the COVID19 pandemic.

In respect of the wider outcomes, including patient satisfaction and regret, the cohort surveyed is biased by only including those who completed testing, with a relatively low response rate (27.3%). We were unable to capture the experiences of people who did not complete testing via the pathway. We also observed differences in demographics in responders compared with the reference cohort.

Finally, whilst HCP survey responses were received from breast and genetics specialities, overall number of responses were small. Furthermore, questions may have been open to interpretation with some survey outputs (such as referrals by different routes) being subjective rather than quantified.

### Conclusions

Our NT-MBGT programme demonstrates feasibility, acceptability, and high satisfaction with a ‘BRCA-DIRECT’ pathway across diverse breast oncology centres in a ‘real world’ setting. Despite the expanded testing eligibility criteria within the programme leading to higher gPV pick-up rates, both breast and genetics clinical services were overall supportive of the pathway. Seemingly reducing impact on clinical time via simplified eligibility assessment, increased mainstreaming, and reduced referrals to clinical genetics. Comprehensive analysis of detection rates enabled exploration of eligibility criteria, balancing simplified eligibility assessment, overall detection (sensitivity), and testing volumes (specificity).

## Data Availability

Patient-level data underlying this article cannot be shared publicly due to data protection requirements. All other data produced in the present study are available upon reasonable request to the authors.

## Disclaimers

This work represents independent research supported by the National Institute for Health and Care Research (NIHR) Biomedical Research Centre at The Royal Marsden NHS Foundation Trust and the Institute of Cancer Research, London. The views expressed are those of the authors and not necessarily those of the NIHR or the Department of Health and Social Care.

