## Supplemental Tables for "Routine germline genetic testing in 3552 unselected NHS breast cancer patients: Evidence informing testing criteria and implementation of a ‘BRCA-DIRECT’ mainstreaming pathway"

**Running head:** ‘BRCA-DIRECT’ mainstreaming pathway

**Authors:** Bethany Torr (MSc)^1,2†^; Lea Mansour (MSc)^3†^; Caitlin Fierheller (PhD)^3^; Monica Hamill (BScN)^1,2^; Joshua Nolan (MSc) ^1,2^; Nicola Bell (BSc) ^1,2^; Subin Choi (MSc)^1^; Sophie Allen (MSc)^1^; Sudeekshna Muralidharan (BSc) ^1,2^; Suzanne MacMahon (PhD)^4^; Yasmin Clinch (MSc)^4^; Mikel Valganon-Petrizan (PhD)^4^; Helena Harder (PhD)^5^; Alice Garrett (MD, PhD)^1,6^, D. Gareth Evans (MD, PhD)^7,8^ Angela George (MD)^1,2^; Valerie Jenkins (DPhil)^5^; Lesley Fallowfield (DPhil)^5^; Rosa Legood (PhD)^9^; Zoe Kemp (MD, PhD)^1,2,11^; Ranjit Manchanda (MD, PhD)^3,9,10*^ , Clare Turnbull (MD, PhD)^1,2*^

*,†: these authors contributed equally to this work

**Affiliations:**

(1) The Division of Genetics and Epidemiology, The Institute of Cancer Research, London, UK
(2) Cancer Genetics Unit, The Royal Marsden NHS Foundation Trust, London, UK
(3) Wolfson Institute of Population Health, Queen Mary University of London, London, UK
(4) Clinical Genetics, The Royal Marsden NHS Foundation Trust, London, UK
(5) Sussex Health Outcomes Research & Education in Cancer (SHORE-C), Brighton and Sussex Medical School, Universities of Brighton and Sussex, Brighton, UK
(6) Department of Clinical Genetics, St Georges Hospital NHS Trust, London, UK
(7) Nightingale and Genesis Breast Cancer Centre, Manchester University Hospitals NHS Foundation Trust, Manchester, UK
(8) Division of Evolution, Infection, and Genomic Sciences, The University of Manchester, Manchester, UK
(9) Department of Health Services Research and Policy, London School of Hygiene and Tropical Medicine, London, UK
(10) Department of Gynaecological Oncology, Barts Health NHS Trust, London, UK
(11) Breast Oncology Unit, The Royal Marsden NHS Foundation Trust, London, UK

Supplementary Table 1 Reference cohort definition

| Inclusion | | - recorded episode of breast cancer (ICD-10 code C50, excluding Stage 0 disease (Paget’s)), or high-grade DCIS (ICD-10 code D051, grades G3, G4, and GH only). - diagnosed between 01-01-2021 and 31-12-2021 - Treating hospital recorded under an NHS trust participating in the programme |
| --- | --- | --- |
| Age | | Recorded age at diagnosis |
| IMD quintile | | Recorded IMD quintile |
| Triple Negative Status | Yes | ER, PR, and HER2 hormone receptor all negative |
|  | No | ER, PR, and HER2 hormone receptors all positive, or mixture of hormone receptor positive and negative |
|  | Unknown | Data from at least one hormone receptor is missing |
| Breast Cancer Status | Primary | Single breast cancer reported (Stage 1-3 or no stage data available), no data on recurrence **OR** Multiple breast cancer reported (Stage 1-3 or no stage data available, no data on recurrence, does not meet definition for bilateral) with first instance of BC within reference year |
|  | Bilateral | Two breast cancers reported, one in left breast and one in right breast (Stage 1-3 or no stage data available), no data on recurrence |
|  | Known recurrence | Breast cancer with date of first recurrence recorded (Stage 1-3 or no stage data available) **OR** Multiple breast cancer reported (Stage 1-3 or no stage data available, no data on recurrence, does not meet definition for bilateral) with first instance of BC prior to reference year |
|  | Metastatic | Breast cancer of stage 4 reported |

Supplementary Table 2: Germline pathogenic variants identified in High Penetrance (HP-) and Intermediate Penetrance (IP-) Breast Cancer Susceptibility Genes(BCSGs) based on pathology adjusted Manchester Score.

| Pathology-adjusted Manchester score | **All genes** | | | **HP-BCSGs** | | **IP-BCSGs** | |
| --- | --- | --- | --- | --- | --- | --- | --- |
|  | n | gPVs identified | Pick up rate | gPVs identified | Pick up rate | gPVs identified | Pick up rate |
| <3 | 984 | 24 | 2.4% | 8 | 0.8% | 16 | 1.6% |
| 3 to 7 | 1376 | 42 | 3.1% | 31 | 2.3% | 11 | 0.8% |
| <7 | 2129 | 55 | 2.6% | 30 | 1.4% | 25 | 1.2% |
| 7 to 11 | 821 | 50 | 6.1% | 38 | 4.6% | 12 | 1.5% |
| 12 to 14 | 251 | 29 | 11.6% | 24 | 9.6% | 5 | 2.0% |
| 15 to 19 | 201 | 17 | 8.5% | 15 | 7.5% | 2 | 1.0% |
| 20+ | 115 | 15 | 13.0% | 15 | 13.0% | 0 | 0.0% |

Supplementary Table 3: Age and IMD quintile distributions in survey respondents compared with the BC reference cohort

| **Category** | | **BC reference cohort** | **Survey Respondents** | | **p-value** |
| --- | --- | --- | --- | --- | --- |
|  |  | **Percent** | **n** | **Percent** |  |
| **Age group** | Less than 40 | 6.47 | 9 | 1.51 | <0.001 |
|  | <49 | 21.79 | 83 | 15.44 |  |
|  | 50-59 | 26.82 | 147 | 24.66 |  |
|  | 60-69 | 23.52 | 172 | 28.86 |  |
|  | 70 and over | 27.87 | 185 | 31.04 |  |
| **IMD quintile** | 1 (Most deprived) | 13.3 | 32 | 5.44 | <0.001 |
|  | 2 | 26.96 | 106 | 18.03 |  |
|  | 3 | 23.82 | 123 | 20.92 |  |
|  | 4 | 19.29 | 140 | 23.81 |  |
|  | 5 (Least deprived) | 16.63 | 187 | 31.8 |  |
|  | Total | 100 | 588 | 100 |  |
